# Reduction in left frontal alpha oscillations by transcranial alternating current stimulation in major depressive disorder is context-dependent in a randomized-clinical trial

**DOI:** 10.1101/2021.06.17.21258764

**Authors:** Justin Riddle, Morgan L. Alexander, Crystal Edler Schiller, David R. Rubinow, Flavio Frohlich

## Abstract

**Background:** Left frontal alpha oscillations are associated with decreased approach motivation and have been proposed as a target for non-invasive brain stimulation for the treatment of depression and anhedonia. Indeed, transcranial alternating current stimulation (tACS) at the alpha frequency reduced left frontal alpha power and was associated with a higher response rate than placebo stimulation in patients with major depressive disorder (MDD) in a recent double-blind placebo controlled clinical trial.

**Methods:** In this current study, we aimed to replicate such successful target engagement by delineating the effects of a single session of bifrontal tACS at the individualized alpha frequency (IAF-tACS) on alpha oscillations in patients with MDD. Electrical brain activity was recorded during rest and while viewing emotionally-salient images before and after stimulation to investigate if the modulation of alpha oscillation by tACS exhibited specificity with regards to valence.

**Results:** In agreement with the previous study of tACS in MDD, we found that a single session of bifrontal IAF-tACS reduced left frontal alpha power during the resting state when compared to placebo. Furthermore, the reduction of left frontal alpha oscillation by tACS was specific for stimuli with positive valence. In contrast, these effects on left frontal alpha power were not found in healthy control participants.

**Conclusion:** Together these results support an important role of tACS in reducing left frontal alpha oscillations as a future treatment for MDD.

**National Clinical Trial:** NCT03449979, “Single Session of tACS in a Depressive Episode (SSDE)” https://www.clinicaltrials.gov/ct2/show/NCT03449979

## 1. Introduction

The left prefrontal cortex is recruited during the processing of positive emotions [1], and this region has been found to be inhibited, indexed by increased alpha frequency power, during the processing of positive emotions in individuals with depression [2] and preclinical dysphoria [3]. Thus, non-invasive brain stimulation interventions for major depressive disorder (MDD) targeted the left frontal cortex with the goal of increasing neural activity (decreasing alpha) [4-6]. However, the first transcranial magnetic stimulation (TMS) protocol that was FDA-approved for the treatment of depression delivered pulse trains to left prefrontal cortex in the alpha frequency (10 Hz) [7, 8]. This approach is counterintuitive because alpha frequency electrical activity is inhibitory to neural activity [9]. Nonetheless, the treatment approach was successful, which led to the theory that amplifying pathological activity (i.e., further increasing alpha power) may cause a homeostatic rebound that results in reduced alpha power after stimulation [10].

We recently performed the first randomized controlled trial of transcranial alternating current stimulation (tACS) in patients with MDD in which we delivered synchronized alpha frequency (10 Hz) current to left and right prefrontal cortex [11]. We found that four consecutive days of stimulation produced a lasting decrease in left frontal alpha power in the eyes-open, resting-state, compared with baseline. These findings are consistent with the interpretation that stimulation disinhibited activity in the left frontal cortex. However, this initial study was conducted with a small sample size (N=18 for relevant conditions) and only in patients with MDD. Despite the pre-registered successful change in the targeted left frontal alpha oscillations, the lack of understanding how and when homeostatic network reorganization occurs make this finding counterintuitive. Thus, to better understand the immediate impact of tACS on left frontal alpha oscillations, we conducted a double-blinded, parallel-arm, placebo-controlled clinical trial in patients with MDD (n=41) and with an age- and sex-matched control group (n=41) in which we delivered 40 minutes of tACS targeted to bilateral frontal cortices and recorded eyes-open, resting-state electroencephalography (EEG) before and after stimulation. The frequency of stimulation was personalized by identifying and targeting the individual alpha frequency (IAF). By matching the stimulation frequency to the peak frequency of endogenous activity for each individual, we hypothesized that the ability for tACS to modulate brain activity would be increased [12]. The primary outcome measure was a reduction in left frontal alpha oscillations with verum stimulation relative to sham for patients with MDD (National Clinical Trial 03449979). In addition, participants passively viewed emotional images from the International Affective Picture System (IAPS) before and after stimulation. Since elevated amplitude of left frontal alpha oscillations is theorized to correspond to a reduction in approach towards positive experiences [1, 2], we hypothesized that stimulation may produce a selective decrease in left frontal alpha oscillations to images rated as positive. The addition of an emotional context provided the ability to investigate whether the impact on left frontal alpha oscillations was context-dependent. Furthermore, the inclusion of a control group allowed us to determine whether tACS effects on left frontal alpha oscillations are specific to depression or whether simulation produces a rebound effect in all participants. Finally, recent evidence indicates that hyperconnectivity in prefrontal cortex may index depression severity [13-17]. In an exploratory functional connectivity analysis, we attempted to replicate this finding that would lend further insight into our strategy of synchronizing the left and right prefrontal cortex with synchronized stimulation.

## 2. Materials and Methods

The experiment was approved by the Institutional Review Board at the University of North Carolina at Chapel Hill. Participants recruited from the Raleigh-Durham-Chapel Hill community provided written consent before participation. The experiment was conducted in the Carolina Center for Neurostimulation from September 2018 to August 2019. The experimental design consisted of two groups, those with MDD and those without MDD (i.e., euthymic “controls”), in which each participant received either tACS or an active sham stimulation in a parallel arm design to improve participant-blinding (see Figure S1, CONSORT Diagram). The experiment was pre-registered on ClinicalTrials.gov where the complete protocol can be found (NCT03449979). First, eyes-closed resting-state EEG was acquired at the start of the experiment. These data were used to localize IAF (Figure 1A), which was used as the frequency for tACS (Figure 1B). Second, participants performed a streamlined version of the Expenditure of Effort for Reward Task; these results are reported in a different manuscript [18]. Third, participants passively viewed images from the IAPS while EEG was acquired. Fourth, eyes-open resting-state EEG was acquired just prior to the start of IAF-tACS. During stimulation, EEG was not collected, as these data are corrupted by the stimulation waveform. Immediately following IAF-tACS, eyes-open resting-state EEG was recorded followed by passive viewing of novel images from the IAPS. Finally, previously viewed emotional images were presented a second time in a random order and participants provided a subjective rating of their emotional reaction to each of the images.

**Figure 1.**
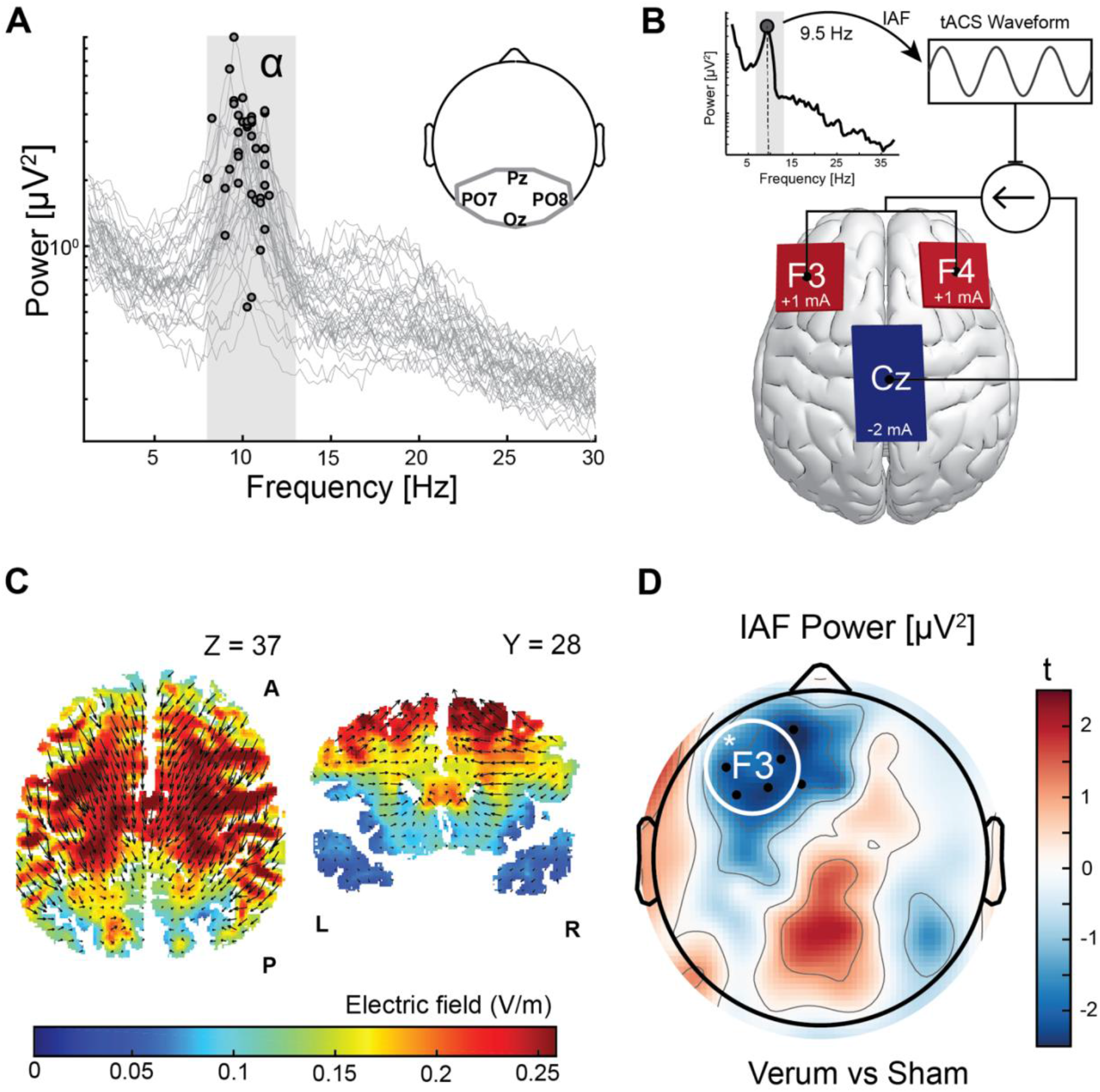
IAF-tACS decreased left frontal IAF power in patients with MDD. (A) Individual alpha frequency (IAF) was localized using an eyes-closed resting-state EEG. The power spectrum from occipital-parietal electrodes (outline of region of interest shown in insert) for each participant is depicted. The frequency with maximum power within the alpha band from 8-13 Hz (labeled grey box) is denoted with a black circle for all patients with MDD. (B) After localizing IAF for each participant, transcranial alternating current stimulation (tACS) was applied to the scalp at IAF. A hypothetical participant is depicted. Dotted line shows the IAF with maximum power in the grey rectangle of the canonical alpha band. IAF-tACS delivered identical current at 1 mA zero-to-peak with a split-wire to two 5×5cm stimulator pads on the left and right frontal cortices (over F3 and F4). The 5×7cm return stimulator pad over Cz acted as an electrical sink. (C) Electric field model shows the regions with maximum electric field strength (V/m) and depicts the electric force lines. On the left, an axial view at the +37 z-plane of Montreal Neurological Institute (MNI) space oriented anterior (A) to posterior (P). On the right, a coronal view at the +28 y-plane of MNI space oriented left (L) to right (R). (D) IAF-tACS produced a selective decrease in IAF power for verum versus sham (modulation index) over the left frontal region of interest (white circle, * p < 0.05). Electrodes with a significant change are depicted with a black dot, p < 0.05, that were in a cluster of at least three contiguous significant electrodes.

### 2.1. Participants and Assessment of Depression Severity

Men and women, ages 18-65, with normal or corrected-to-normal vision were recruited using ads in the community. Potential participants completed a brief phone screening, and 126 participants were enrolled in the experiment (CONSORT Diagram). After applying inclusion and exclusion criteria (Supplemental Section 1), 87 participants were randomized to receive either IAF-tACS or sham-tACS and 82 participants were included in the final analysis (66 women): 41 healthy controls and 41 patients with MDD. Our analyzed sample size goal was at least 80 participants, which was determined to exceed 1 – beta > 0.95 based on the large effect-size on left frontal alpha power in our previous clinical trial with this stimulation montage in the same study population [11]. In each group, 21 participants received verum stimulation (17 women) and 20 participants received an active sham (16 women). Depression severity was quantified using the Hamilton Depression Rating Scale (HAM-D) and Beck Depression Inventory version 2 (BDI-II), and was used in individual differences analyses. Descriptive statistics for the patients with MDD and healthy control population are reported in Table 1 and demographic information for patients with MDD is reported in Table 2.

**Table 1.**
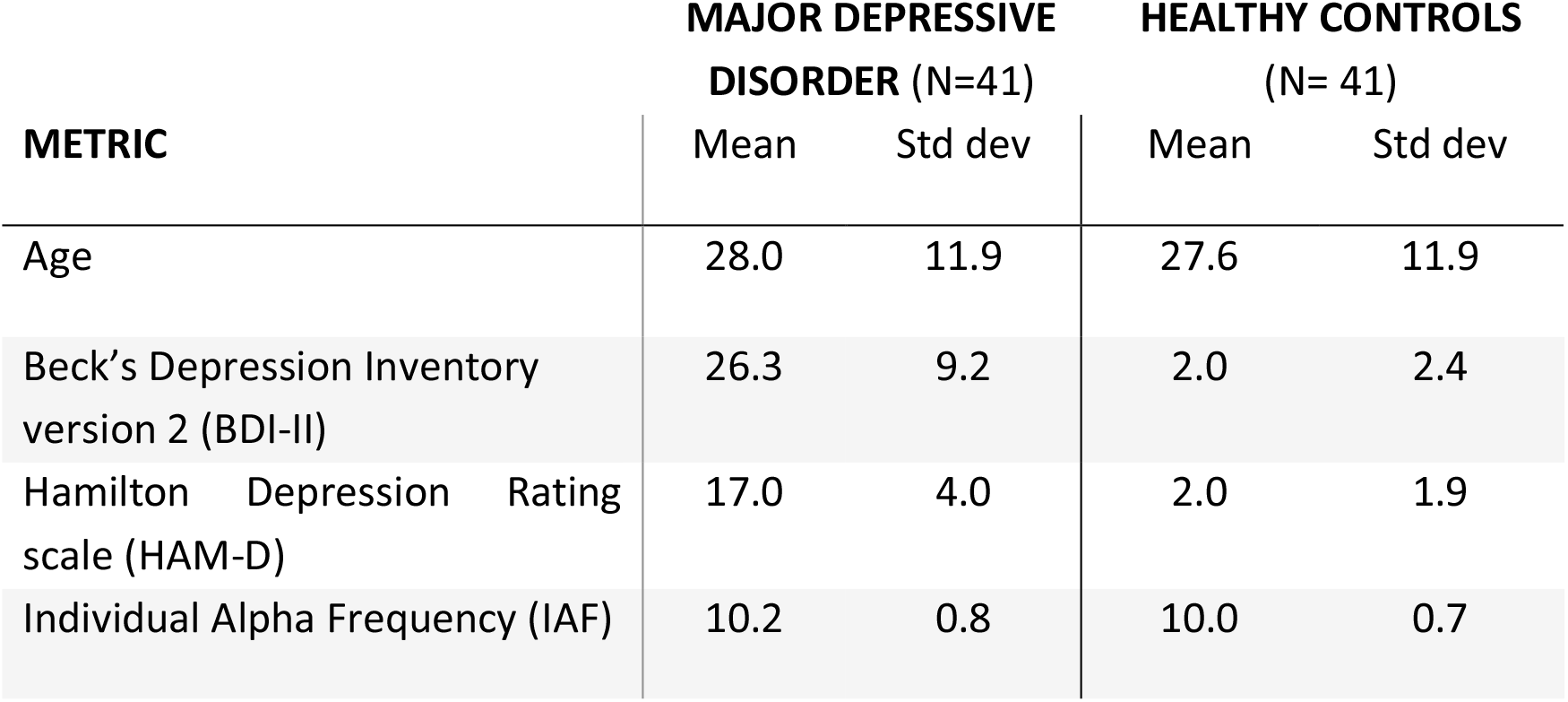
Descriptive statistics for patients with MDD and for age and sex-matched healthy controls. Individual alpha frequency (IAF) was calculated with minimal preprocessing during the experiment from parietal-occipital electrodes. The resolution of IAF was 0.5 Hz resolution. IAF was used for stimulation and for subsequent analysis of stimulation effects on resting-state EEG data.

| METRIC | MAJOR DEPRESSIVE<br>DISORDER (N=41) |  | HEALTHY CONTROLS<br>(N= 41) |  |
| --- | --- | --- | --- | --- |
|  | Mean | Std dev | Mean | Std dev |
| Age | 28.0 | 11.9 | 27.6 | 11.9 |
| Beck's Depression Inventory<br>version 2 (BDI-II) | 26.3 | 9.2 | 2.0 | 2.4 |
| Hamilton Depression Rating<br>scale (HAM-D) | 17.0 | 4.0 | 2.0 | 1.9 |
| Individual Alpha Frequency (IAF) | 10.2 | 0.8 | 10.0 | 0.7 |

**Table 2.** Descriptive information specific to patients with major depressive disorder. * for education level 1-5 is some high school, high school, some college or associates degree, bachelor’s degree, advanced degree. ^ median and median absolute deviation used due to outliers. SSRI = selective serotonin reuptake inhibitor. NDRI = norepinephrine-dopamine reuptake inhibitor. “Other” includes buspirone, trazodone, brexpiprazole, amitriptyline.

| MDD (N = 41) | VERUM (N = 21) | SHAM (N = 20) |
| --- | --- | --- |
| Beck's Depression Inventory version 2<br>(BDI-II) | 25.09 (9.20) | 27.55 (9.25) |
| Hamilton Depression Rating scale<br>(HAM-D) | 16.43 (4.19) | 17.65 (3.79) |
| Education* | 2.67 (0.97) | 2.50 (0.83) |
| Depression Occurs in Episodes? | 15 / 21 | 17 / 20 |
| - Duration Current Episode (months)^ | 3.00 (2.93) | 2.00 (13.95) |
| Maudsley Staging Method | 6.28 (2.08) | 6.25 (1.97) |
| Medication for Mood Disorder? (last<br>year) | 10 / 21 | 12 / 21 |
| Medication for Mood Disorder? (last two weeks) | 8 / 21 | 9 / 21 |
| - Current Medication (SSRI) | 7 / 7 | 6 / 9 |
| - Current Medication (NDRI) | 3 / 7 | 2 / 9 |
| - Current Medication (Other) | 1 / 7 | 3 / 9 |
| Psychotherapy (last year) | 15 / 21 | 10 / 20 |
| Psychotherapy (last two weeks) | 7 / 21 | 6 / 20 |
| Comorbid Anxiety Disorder | 12 / 21 | 11 / 20 |
| Smoke tobacco? (Yes, how many cigarettes per week?) | 1 / 21 (5.00) | 2 / 20 (0.75 avg) |
| Drink alcohol? (Yes, how many drinks per week?) | 11 / 21 (2.81 avg) | 15 / 20 (2.57 avg) |

### 2.2. Preprocessing of the Electroencephalogram

EEG data were collected with a high-density 128-channel electrode net at 1000 Hz (HydroCel Geodesic Sensor Net) and EGI system (NetAmps 410, Electrical Geodesics Inc., OR, USA). The impedance of each electrode was below 50 kΩ at the start of each session. Because transcranial alternating current stimulation (tACS) was delivered using three conductive electrodes of sizes five by five centimeters to F3 and F4 and five by seven centimeters to Cz, electrodes around these regions were bridged. The Cz electrode which served as the reference electrode in the EGI system was placed directly on the scalp via a hole cut in the stimulation electrode surrounding Cz. A typical preprocessing pipeline was used for the EEG data during resting-state and emotional image viewing (e.g., [19, 20]) (Supplemental Section 2).

### 2.3. Transcranial Alternating Current Stimulation

The electrode montage for tACS was adapted from our previous randomized clinical trial of MDD [11]. However, in our previous experiment, stimulation was delivered at a fixed frequency in the center of the canonical alpha band (10 Hz). In the current experiment, stimulation was delivered at the IAF, because previous work has demonstrated that stimulation is able to more effectively entrain neural activity to the stimulation waveform when it is matched to the frequency of the targeted neural activity [12, 21]. To localize IAF, we recorded two minutes of eyes-closed resting-state EEG before any other recordings. These data were analyzed during the acquisition of task data (Figure S1) using an abbreviated preprocessing pipeline (Supplemental Section 3) and used for stimulation and subsequent analysis of resting-state EEG data. IAF was derived from parietal-occipital electrodes and this frequency was comparable to IAF derived from left frontal electrodes (Figure S2). IAF-tACS was delivered using a lightweight battery-powered device with a paired tablet that was designed for double-blinded clinical trials (XCSITE 100, Pulvinar Neuro LLC, Chapel Hill, NC, USA) using the same protocol as in our previous study [11] (Supplemental Section 4). The induced electric field from stimulation was estimated using the ROAST toolbox estimated on a template brain [22]. Electric field models show that current flowed mostly in the anterior-posterior orientation (Figure 1C).

### 2.4. Spectral Analysis

Spectral analysis was run on the four-minute eyes-open resting-state EEG data before and after stimulation using the fast Fourier transform on every two-second epoch with no overlap. Median power spectra was calculated, and IAF plus or minus 1 Hz was averaged for each channel. IAF power was normalized within each participant by dividing each channel by the sum of the 90 channels on the scalp. In our previous experiment, alpha power in left frontal electrodes showed the strongest modulation from stimulation. Thus, our primary analyses were performed on left frontal electrodes (F3 and the five surrounding electrodes) to reduce multiple comparisons in a pre-registered analysis. An exploratory search of the scalp was performed to understand the spatial-specificity of significant effects. The change in IAF power from IAF-tACS was calculated using the modulation index: 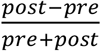 to normalize for differences in the magnitude of IAF power between participants.

### 2.5. Functional Connectivity

In an exploratory analysis, functional connectivity analysis was run for IAF with a seed over left frontal cortex (F3) and a target in an a priori region of interest of right frontal electrodes (F4 and five surrounding electrodes) based on previous findings [16, 17]. Functional connectivity was calculated using weighted phase lag index (wPLI). Channels surrounding F3 – lateral to Fz and anterior to C3 – were removed from the analysis, as connectivity with these channels exceed the spatial resolution of EEG. The data for each recording was normalized by dividing each channel by the sum of all data channels, and the effect of stimulation was normalized using the modulation index.

### 2.5. Emotional Valence Task

Before and after stimulation, participants viewed emotionally salient images from the IAPS [23]. In each of these session, 96 images were presented with an equal number of each valence randomly interleaved (positive, neutral, and negative). Images were presented at fixation with a height of 10 degrees visual angle. Participants were instructed to maintain fixation during image presentation and to blink during the inter-trial interval. Each image was presented for exactly 2 seconds and was submitted to time-frequency analysis. At the end of the experiment, participants were presented with the same 192 images in a random order and were asked to rate each image on a scale from 1-10, utilizing the full scale, on how negative to positive the participant experienced the image (1=very negative, 10=very positive). The image-rating session was self-paced and its EEG data were not analyzed.

We were interested in alpha power in the left frontal cortex during image viewing as a function of valence. Trials were categorized based on subjective rating collected at the end of the experiment. Positive images were rated 8 to 10 (number of trials was 39.5±21.9), neutral was 4 to 7 (93.3±34.1), and negative was 1 to 3 (59.2±19.9). Two of the 41 patients with MDD did not have usable data due to technical errors. Four patients with MDD rated less than five images as positive. Thus, analyses for the positive condition comprised 18 patients with MDD that received verum and 17 that received sham. Time-frequency analysis was run by convolving trial data (mirrored to reduce edge artifacts) with five-cycle Morlet wavelets of 150 frequencies from 2 to 59 Hz spaced along an adjusted log scale (**Equation 1**).

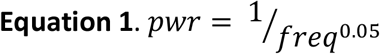

This distribution approximated the naturally-occurring power distribution of human brain activity [24]. Data were averaged for each condition and baseline-corrected in time-frequency domain to −800 to −500 milliseconds from image onset.

### 2.6. Statistical Analysis

To test for successful double-blinding, Chi-square tests of independence were run with stimulation type as group (verum or sham) and the guess made by the participant or experimenter as category. The alpha threshold was set at 0.05 with two-tails for all statistical analyses. Our pre-registered primary outcome was an interaction between time (before or after stimulation) and IAF-tACS (verum or sham) in the patients with MDD. Our follow-up analysis utilized the modulation index to normalize differences in magnitude of left frontal alpha oscillations across patients. Multiple comparisons were reduced by pre-registering our primary outcome, focusing on a single region of interest, defining a single time-frequency cluster from all trials in the emotional valence task (permutation-based cluster correction for mass [25]), and focusing on positive valence [3]. Exploratory analyses include topographic analysis with a significance threshold of three significant contiguous electrodes, individual differences analysis using depression severity, IAF-tACS effect as a function of antidepressant use, the relationship of IAF functional connectivity between left and right prefrontal cortex to depression severity, and the impact of IAF-tACS in healthy control participants.

## 3. Results

### 3.1. Electrical stimulation modulates left frontal alpha power in MDD

Participants were successfully blinded to the stimulation (N=80, two participants did not answer, χ^2^=0.2631, p=0.608). Of the 42 participants that received verum stimulation, 33 believed they received verum stimulation. However, of the 40 participants that received sham stimulation, 28 believed they received verum stimulation (2 chose not to answer). Thus, participants, irrespective of stimulation type, were biased towards believing they received verum and their accuracy was 53.8%, near chance levels. The experimenters were also blind to stimulation (N=82, χ^2^=0.038, p=0.845): accurate guessing for verum stimulation was 42.9% and 55.0% for sham stimulation. Thus, our double-blinding procedure was successful. Adverse events were common side effects and were found in 28.6% of participants for sham stimulation and 50% for verum stimulation.

Our primary outcome was an interaction between time and stimulation for patients with MDD, which was found to have a trend-level effect (F(1,39)=3.119, p=0.0852). By using the modulation index to normalize for differences in magnitude of the alpha oscillation across patients, we found that left frontal IAF power was significantly decreased for verum relative to sham stimulation in patients with MDD (N=41, −0.016±0.007; t(39)=-2.133, p=0.039, d=0.667) (Figure 1D). Post-hoc t-tests on the impact of IAF-tACS on left frontal IAF power for verum and sham stimulation revealed a significant increase in IAF power with sham in patients with MDD (0.012±0.024; t(19)=2.191, p=0.041, d=0.489) (Figure 2A), and a not significant decrease in IAF power with verum (−0.004±0.024; t(20)=-0.802, p=0.432, d=0.175) (Figure 2B). These results suggest that without verum stimulation left frontal IAF power was increased, but this rise in power was negated by IAF-tACS.

**Figure 2.**
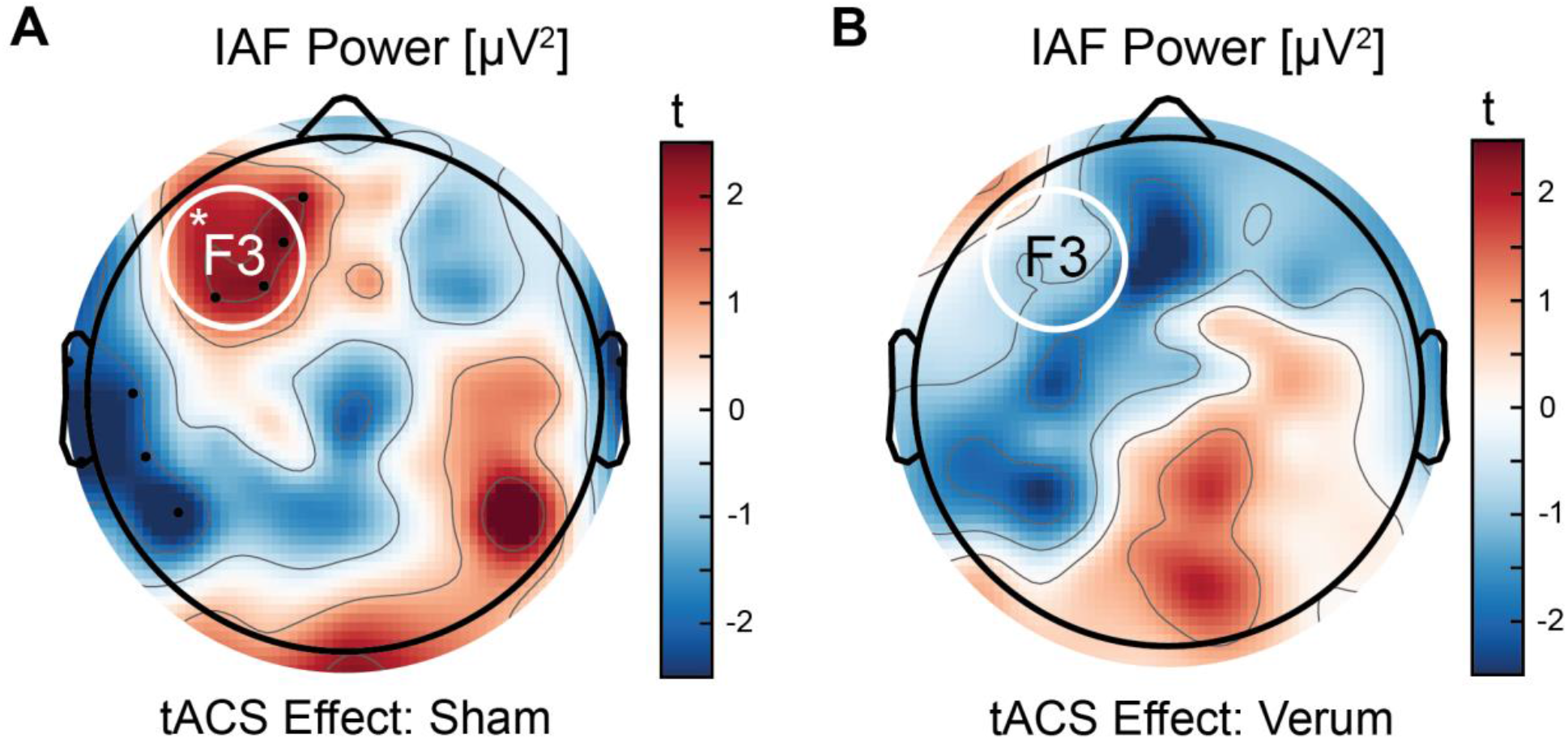
IAF-tACS negated the increase in left frontal IAF power during the resting-state. (A) In the absence of stimulation, a selective increase in IAF power over the left frontal region of interest (white circle, * p < 0.05) was observed. (B) Verum IAF-tACS did not produce a significant change in left frontal IAF power. Electrodes with a significant difference from post- to pre-stimulation are depicted with a black dot, p < 0.05, that were in a cluster of at least three contiguous significant electrodes.

### 3.2. Individual differences in the effect of tACS

We investigated whether greater depression severity in patients with MDD resulted in a stronger modulation of left frontal IAF power from IAF-tACS. Indeed, self-report (BDI-II) correlated with modulation of left frontal IAF power (Fisher’s Z, difference in correlation, z(39)=2.185, p=0.029) (Figure 3A). Similar to the mean effect, post-hoc analysis found a significant relationship for sham, but not verum, stimulation (Figure S3). However, we did not find a significant relationship using a clinician-administered assessment (HAM-D) (z(39)=1.405, p=0.160) (Figure 3B). An additional variable that might mediate the impact of IAF-tACS on left frontal alpha power is medication status. An exploratory analysis found that the impact of IAF-tACS on left frontal alpha power was strongest in patients using an antidepressant (Figure 4).

**Figure 3.**
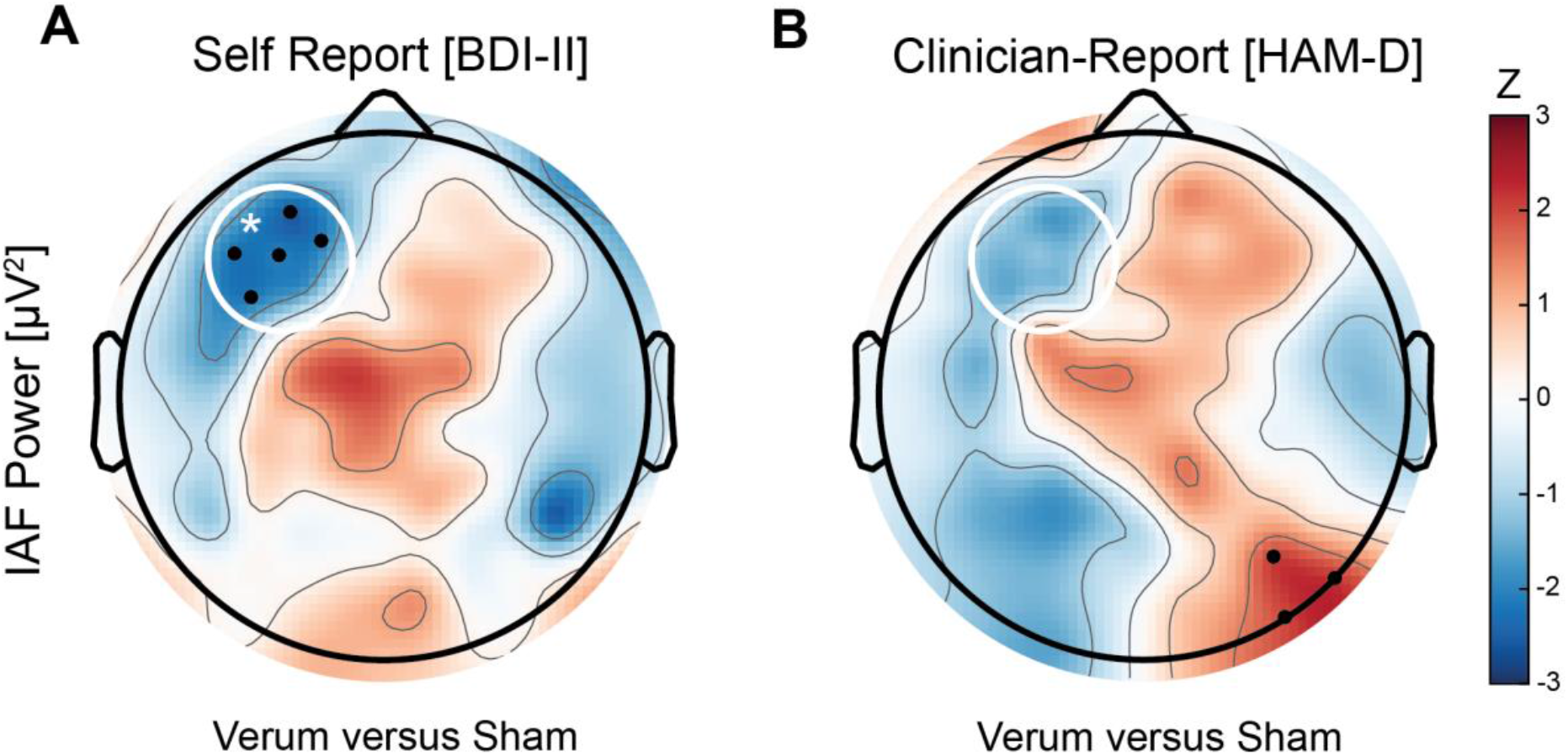
Individual difference in tACS effect by depression severity. Individual differences in the impact of stimulation on IAF power across the scalp was correlated with baseline depression severity in patients with MDD quantified by self-report using the BDI-II (A) and clinician-report using the HAM-D (B). The difference in correlation between the verum and sham group found a focal difference in left frontal electrodes. Left frontal region of interest highlighted with a white circle. * p < 0.05. Dots represent electrodes with a significant difference and at least three contiguous significant electrodes.

**Figure 4.**
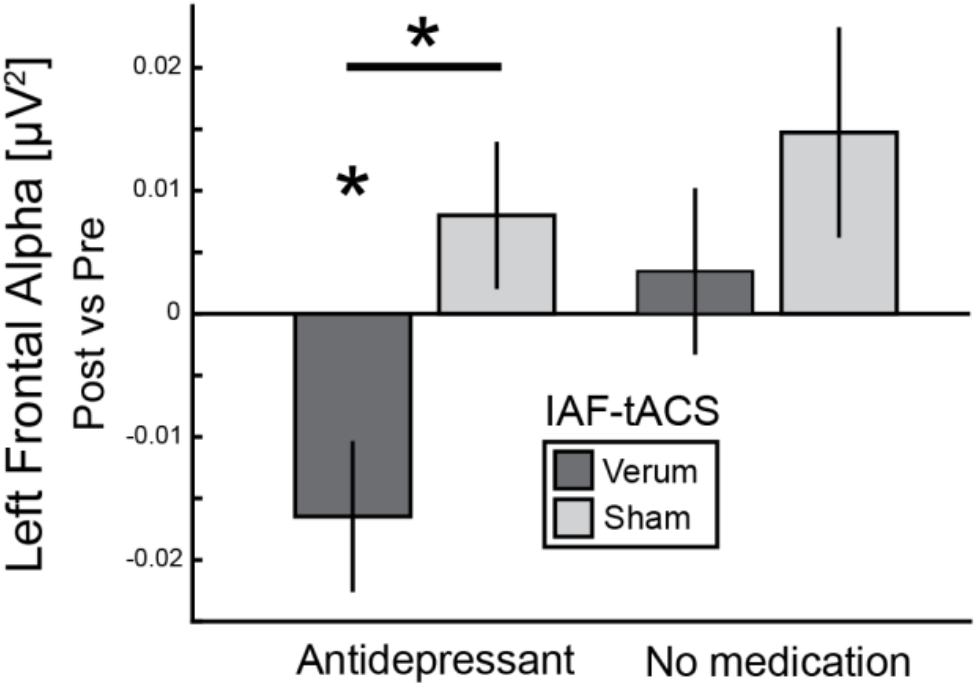
Analysis of the effect of IAF-tACS on left frontal IAF power by medication status. Patients with MDD were categorized by use of an antidepressant within the past two weeks. A two-way ANOVA using between antidepressant-use and stimulation (verum or sham) found a main effect of stimulation (F(1,37)=4.791, p=0.035, η_p_^2^=0.11), a trend-level effect of antidepressant-use (F(1,37)=3.277, p=0.078, η_p_^2^=0.08), and no interaction (F(1,37)=0.801, p=0.377, η_p_^2^=0.02). Post-hoc testing revealed a significant effect of stimulation with antidepressant-use (N=17, t(15)=-2.848, p=0.012, d=1.386) driven by a decrease in left frontal IAF power for verum (N=8, t(7)=-2.683, p=0.031, d=0.949). Without antidepressants there was no significant effect of stimulation (N=24, t(22)=-1.049, p=0.305, d=0.428). Critically, there was no difference in depression severity (HAM-D) between patients groups by antidepressant-use (t(39)=0.124, p=0.902, d=0.0398). *p<0.05; error bars are SEM; units are modulation index.

### 3.3. Stimulation disinhibits left frontal cortex with positive images

Before and after IAF-tACS, participants passively viewed positive, neutral, and negative images (Figure 5A). Across all valences, we found a robust decrease in alpha amplitude from 0.1 to 1.5 seconds after stimulus onset (Figure 5B) used in subsequent analyses (0.1-1.5 seconds; 8-12 Hz). As hypothesized, we found that the amplitude of left frontal alpha oscillations was elevated for positive relative to neutral images at baseline in patients with MDD (N=35, 0.094±0.210; t(34)=2.663, p=0.012, d=0.450) (Figure 5C). There was no significant difference for negative relative to neutral images in left frontal electrodes (N=39, 0.055±0.194; t(38)=1.779, p=0.083, d=0.301) (Figure 5D).

**Figure 5.**
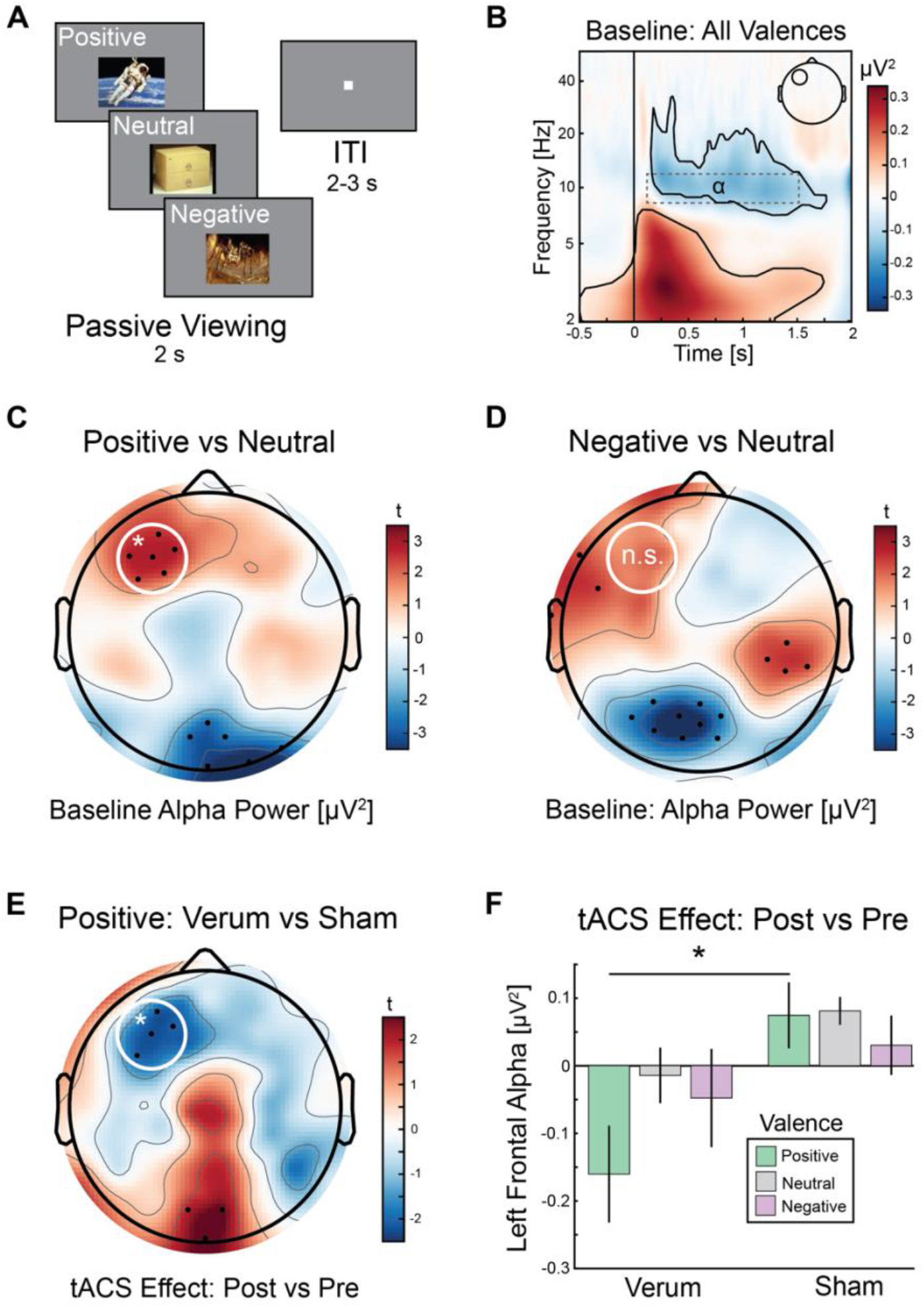
Elevated left frontal alpha power to images rated as positive is inhibited by IAF-tACS in patients with MDD. (A) Patients with MDD passively viewed positive, neutral, and negative images from the IAPS (example images provided). Stimuli were presented for two seconds (s) separated by a two to three second inter-trial interval (ITI). (B) Across all valences at baseline in patients with MDD, time-frequency analysis revealed a significant decrease in alpha amplitude and increase in theta amplitude in left frontal electrodes (insert in the upper right). Black outline shows significant cluster at p < 0.05 after permutation-based cluster-correction by mass. Vertical line at zero denotes onset of the image. The dashed grey rectangle from 0.1 to 1.5 seconds and 8 to 12 Hz is the alpha power that was extracted. (C) At baseline, the contrast of positive versus neutral alpha power revealed a selective increase in left frontal electrodes and decrease in parietal-occipital electrodes. The white outline is the left frontal region of interest. * p < 0.05 for a priori analysis. Black dots represent electrodes with a significant difference that were in a cluster of at least three contiguous electrodes. (D) At baseline, the contrast of negative versus neutral alpha power revealed no modulation in the left frontal region of interest (white circle). “n.s.” is not significant. (E) For the contrast of verum versus sham, there was a selective reduction of left frontal alpha power for positive images. The tACS effect is the change in alpha amplitude for image viewing data post minus pre stimulation. (F) For the left frontal region of interest, the change in alpha power from stimulation is shown for all valence and stimulation conditions. The comparison of interest from the baseline analysis revealed a significant difference for verum versus sham. * p < 0.05. Error bars are within-participant SEM. Alpha power was z-transformed across scalp electrodes so units are z.

With IAF-tACS, we found a significant reduction in left frontal alpha power for positive images relative to sham in patients with MDD (N=35, −0.235±0.113; t(33)=-2.045, p=0.049, d=0.715) (Figure 5E), but no change in subjective ratings for positive images (N=41, verum:6.700±0.780, sham:7.020±1.055, t(38)=-1.089, p=0.283, d=0.348). Post-hoc analysis of the impact of verum and sham stimulation separately revealed a non-significant, decrease in alpha amplitude for verum (N=18, −0.160±0.405; t(17)=-1.678, p=0.112, d=0.396), and no significant change for sham (N=17, 0.075±0.252; t(16)=1.221, p=0.240, d=0.296) (Figure 5F). While stimulation resulted in a decrease in left frontal alpha oscillations during eyes-open resting-state, the decrease in left frontal alpha oscillations during the emotional valence task was specific to images rated as positive and was unchanged for images rated as negative or neutral (Figure S4).

### 3.4. Healthy Controls Did Not Exhibit Left Frontal Alpha Modulation

To better understand the impact of IAF-tACS, we investigated whether healthy control participants exhibited the same increase in left frontal alpha power to positive imagery and to placebo stimulation. At baseline, healthy control participants did not exhibit an increase in left frontal alpha power to positive versus neutral images (two participants did not have sufficient trials, N=39, 0.014±0.235, t(38)=0.379, p=0.707, d=0.061) (Figure 6A). Furthermore, healthy controls did not show differential modulation of left frontal alpha power to positive images between verum and sham (N=38, 0.001±0.113, t(36)=0.013, p=0.990), nor a reduction from verum stimulation (N=19, 0.025±0.296, t(18)=0.369, p=0.716, d=0.085) (Figure 6B). Furthermore, there was no change in left frontal IAF power during eyes-open resting-state from stimulation (time by stimulation, F(1,39)=0.187, p=0.6676). Post-hoc analyses did not a significant difference in left frontal IAF power between verum and sham stimulation (N=41, −0.003±0.007, t(39)=-0.423, p=0.675) (Figure 6C), nor a significant increase from sham stimulation as in patients (N=20, 0.006±0.025, t(19)=1.109, p=0.281, d=0.248) (Figure 6D). Comprehensive ANOVAs performed across all participants for the eyes-open resting-state recapitulated the pattern of findings revealed from our pre-registered hypothesis-based statistical approach (Supplemental Section 5).

**Figure 6.**
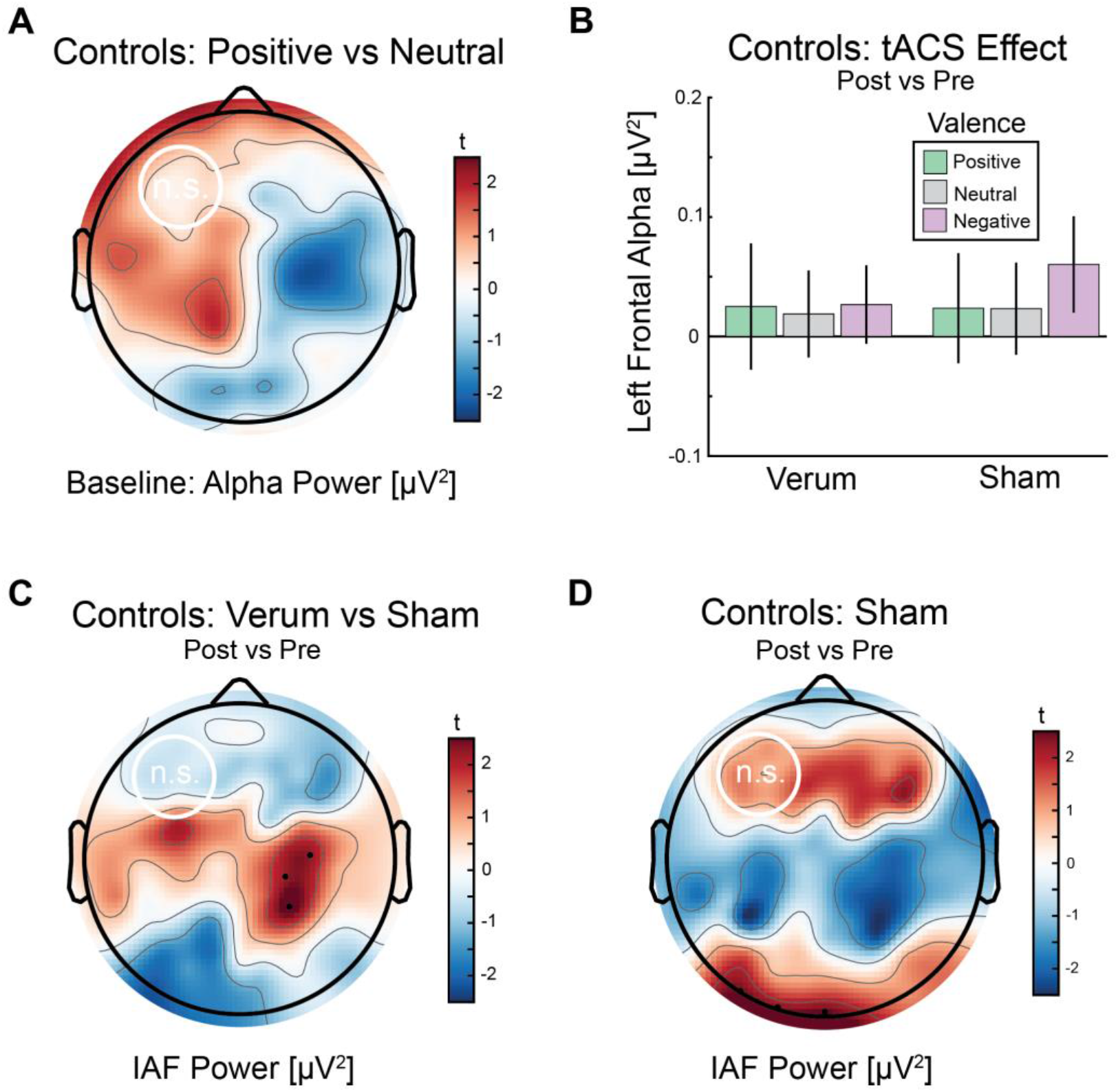
Healthy controls without left frontal alpha engagement show no effect of stimulation. (A) At baseline, there was no increase in left frontal alpha power for positive relative to neutral images. White circle depicts left frontal region of interest. N.s. means not significant. (B) Left frontal alpha power does not show modulation from IAF-tACS in healthy control participants. Error bars are within-participant SEM. (A,B) Units are z-transformed alpha power values across scalp channels. (C) Control participants do not exhibit an impact of IAF-tACS for verum versus sham in left frontal IAF power. (D) Unlike the patients with MDD, healthy controls do not show an increase in left frontal IAF power during resting-state after sham stimulation, as in patients with MDD. Black dots depict p < 0.05 with at least 3 contiguous electrodes. (C,D) Units are modulation index.

### 3.5. Functional connectivity, not power, as a predictor of depression severity

As an exploratory analysis, we investigated a possible relationship between functional connectivity in IAF between left and right frontal cortex and depression severity (HAM-D) in patients with MDD (baseline: 0.012±0.010) and found a significant positive association (N=41, r(40)=0.353, p=0.024) (Figure 7A). By comparison, there was no significant relationship between left frontal IAF power (baseline: 0.009±0.001) and depression severity (r(40)=-0.177, p=0.268) (Figure 7B). Unsurprisingly, there was no significant relationship between depression severity and IAF functional connectivity in healthy controls (N=41, r(40)=-0.025, p=0.874) as there was low variance in depression scores.

**Figure 7.**
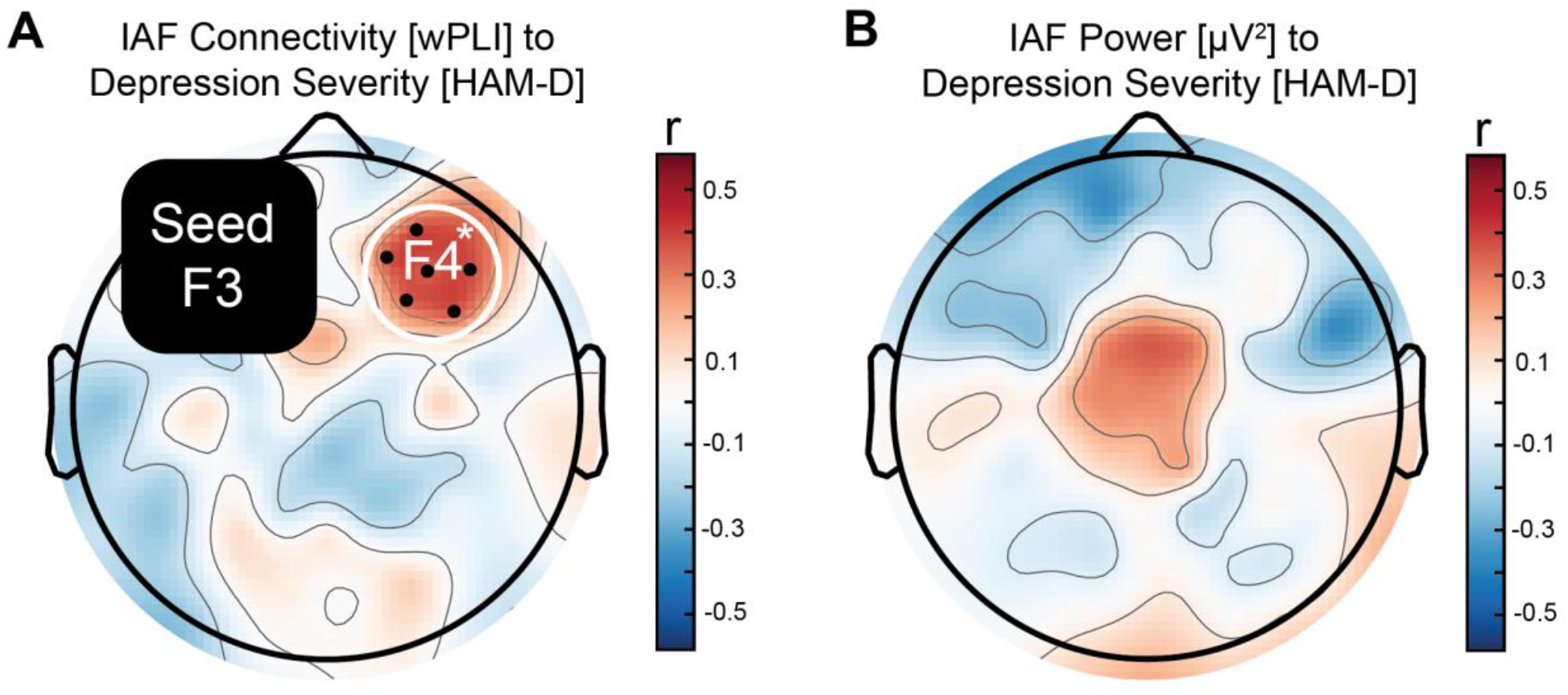
Individual difference in baseline functional connectivity positively tracked with depression severity. (A) Individual differences analysis revealed that depression severity (HAM-D) was positively correlated with functional connectivity strength (weighted phase lag index, wPLI) between the left and right frontal cortices in patients with MDD. The artifact zone surrounding the seed in F3 is depicted with a black box with rounded corners. A dot here denotes an electrode with a correlation of p < 0.05 and with at least 3 contiguous significant electrodes. A white outline was drawn around F4 and the surrounding electrodes. (B) Individual differences analysis for IAF power did not reveal any significant relationship with depression severity in patients with MDD. Functional connectivity and IAF power values were divided by the sum of scalp channels.

## 4. Discussion

Patients with MDD were recruited to receive 40-minutes of transcranial alternating current stimulation (tACS) in the individual alpha frequency (IAF) designed to reduce left frontal alpha oscillations; electrophysiology was recorded before and after stimulation. In agreement with our previous clinical trial that applied five consecutive days of tACS in patients with depression [11], we found a selective decrease in left frontal IAF power for verum versus sham stimulation. The effect was driven by an increase in left frontal IAF power for sham stimulation that was negated by verum stimulation. Individual difference analysis demonstrated that the impact of stimulation of tACS on left frontal IAF power scaled with greater depression severity as measured by self-report. Furthermore, the effects were strongest in those taking antidepressants within two weeks of IAF-tACS. To understand the context-dependent nature of elevated left frontal alpha oscillations, participants viewed positive, neutral, and negative images from the International Affective Picture System (IAPS). At baseline, patients with MDD showed elevated left frontal alpha response to positive images relative to neutral images. As a function of stimulation (verum versus sham), patients with MDD showed a reduction in left frontal alpha power in response to positive images. Together, these findings suggest that left frontal alpha power inversely tracks with approach toward positive experiences. Healthy controls failed to show an increase in left frontal alpha either to positive images or during resting-state with sham stimulation; and, thus, there was no effect of stimulation as there was no oscillopathy to negate. As an exploratory analysis, clinician-rated depression severity positively correlated with IAF functional connectivity strength between the left and right frontal cortices. Altogether, stimulation that delivers synchronized current to bilateral prefrontal cortex may disinhibit the left frontal cortex to approach positive experiences in patients with MDD.

The finding that verum stimulation did not have any impact on left frontal alpha in the presence of negative or neutral images suggests the importance of assessing cognitive and emotional state. Our previous clinical trial relied solely on an analysis of resting-state data. While the impact of stimulation was discernible and replicated in this study, the interpretation for how that activity relates to cognition or experience was inherently limited. By including conditions with emotionally salient images, the dynamic reaction of the brain was highly informative, as positive, but not negative or neutral, images resembled the eyes-open resting-state. These findings suggest that the affective state of the participant is critical. Thus, systematic differences between laboratory environments could theoretically alter neural activity during resting-state, which may explain failures to replicate for alpha-frequency power-based analyses.

Hypoactivation of the left prefrontal cortex has been a rather consistent finding in depression research, often associated with behavioral activation, or the pursuit of experience that are deemed to be rewarding [26-31]. Investigations with patients with MDD performing reward-based decision-making tasks have consistently found impairments in reward learning and motivation [32-34]. These deficits may be dependent on impaired left prefrontal cortical activation [32], in which case IAF-tACS could be applied to improve reward learning or goal-directed behavior that in turn would alleviate symptoms of depression [35]. Combining non-invasive brain stimulation with psychotherapeutic interventions that foster the pursuit of value-based, positive activities such as behavioral activation may be of clinical utility [36].

As with any scientific investigation, the present study has limitations. First, we did not measure levels of subjective arousal to the IAPS images. Thus, we cannot rule out a systematic difference in arousal for the positive versus the neutral and negative images. However, positive and negative images tend to evoke higher arousal than neutral images [37]. Thus, the specificity of our stimulation effects to the positive imagery is not likely explained by arousal differences. Second, we could not record electrophysiology during stimulation [38, 39], so we cannot confirm whether differences in neural entrainment to the stimulation waveform drove our effects. Finally, our experiment was not designed to systematically investigate additional important factors that might mediate the efficacy of stimulation such as medication class, treatment resistance, or demographic factors.

These findings are consistent with the homeostatic theory for targeting oscillopathy [10], in which the pathology is increased with stimulation in order to engage a biological reset. Future work is required to establish what qualifies as a maladaptive oscillation, leading to an oscillopathy, and whether some inherent quality makes this type of neural activity susceptible to homeostatic reset. This model is challenged by the fundamental difficulty in defining what is maladaptive, as the argument could be made that the system is precisely optimized, but towards a maladaptive goal-state. An alternative model is that stimulation delivered in an inhibitory band (e.g. alpha or beta band) drives a network reconfiguration, whereas stimulation delivered in an excitatory band (e.g. theta or gamma) does not. A recent experiment that delivered stimulation targeted to inhibitory activity produced a decrease in behavioral performance and reconfigured the targeted network, whereas targeting excitatory activity improved performance and enhanced network activity [19]. Furthermore, stimulation in inhibitory bands may disrupt cognitive processing during stimulation, but might confer cognitive or mood benefits after network reconfiguration. For example, beta-frequency (20 Hz) stimulation to the hippocampal memory network during task performance caused disruption of associative memory [40], but five consecutive days of stimulation in beta-frequency produced a lasting improvement in associative memory when tested in a follow-up session [41]. Future research should investigate the potential for non-invasive brain stimulation to disrupt maladaptive network activity by delivering stimulation in a frequency associated with neuronal inhibition, and subsequent sessions to consolidate network changes [42].

## Data Availability

The data is available upon request.

## Acknowledgements

This study was supported in part by the National Institute of Mental Health of the National Institutes of Health under Award Numbers R01MH101547 and R01MH111889 awarded to FF and the postdoctoral training program (JR) in reproductive mood disorders T32MH09331502 awarded to DR. Thanks to Alana K. Atkins and Trevor McPherson for help with data collection. Thanks to Trevor McPherson for assistance with coding the adjusted log-distribution and experimental presentation scripts. Thanks to Regina Lapate for consultation early in project development.

---

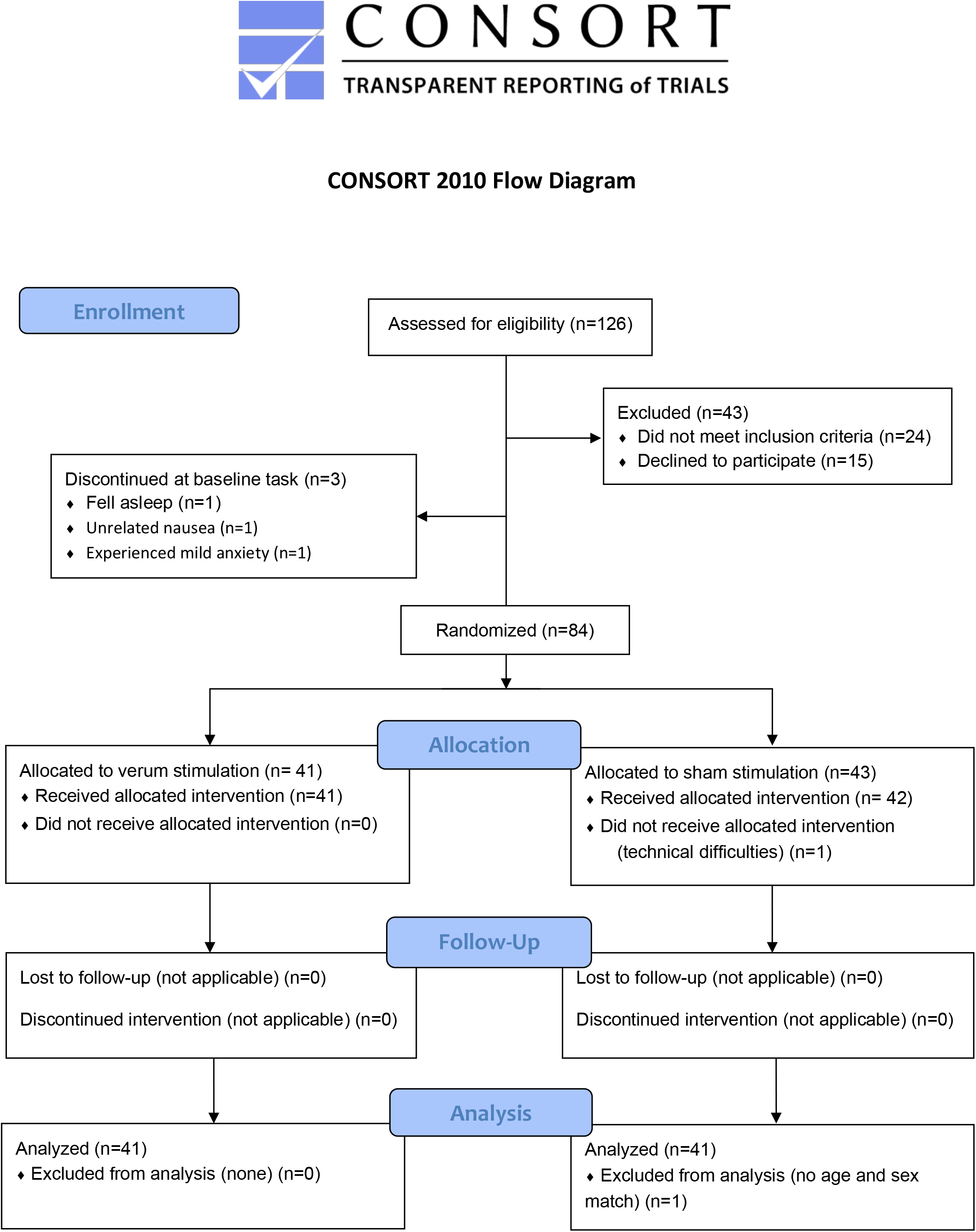

## References

1. Davidson, R.J., Anterior cerebral asymmetry and the nature of emotion. Brain and cognition, 1992. 20(1): p. 125–151.

2. Harmon‐Jones, E. and P.A. Gable, On the role of asymmetric frontal cortical activity in approach and withdrawal motivation: An updated review of the evidence. Psychophysiology, 2018. 55(1): p. e12879.

3. Benvenuti, S.M., et al., Appetitive and aversive motivation in depression: The temporal dynamics of task-elicited asymmetries in alpha oscillations. Scientific Reports, 2019. 9(1): p. 1–11.

4. Allen, J.J., et al., The stability of resting frontal electroencephalographic asymmetry in depression. Psychophysiology, 2004. 41(2): p. 269–280.

5. Pascual-Leone, A., et al., Rapid-rate transcranial magnetic stimulation of left dorsolateral prefrontal cortex in drug-resistant depression. The Lancet, 1996. 348(9022): p. 233–237.

6. Grimm, S., et al., Imbalance between left and right dorsolateral prefrontal cortex in major depression is linked to negative emotional judgment: an fMRI study in severe major depressive disorder. Biological psychiatry, 2008. 63(4): p. 369–376.

7. Perera, T., et al., The clinical TMS society consensus review and treatment recommendations for TMS therapy for major depressive disorder. Brain stimulation, 2016. 9(3): p. 336–346.

8. O’Reardon, J.P., et al., Efficacy and safety of transcranial magnetic stimulation in the acute treatment of major depression: a multisite randomized controlled trial. Biological psychiatry, 2007. 62(11): p. 1208–1216.

9. Goldman, R.I., et al., Simultaneous EEG and fMRI of the alpha rhythm. Neuroreport, 2002. 13(18): p. 2487.

10. Leuchter, A.F., et al., The relationship between brain oscillatory activity and therapeutic effectiveness of transcranial magnetic stimulation in the treatment of major depressive disorder. Frontiers in human neuroscience, 2013. 7: p. 37.

11. Alexander, M.L., et al., Double-blind, randomized pilot clinical trial targeting alpha oscillations with transcranial alternating current stimulation (tACS) for the treatment of major depressive disorder (MDD). Translational psychiatry, 2019. 9(1): p. 1–12.

12. Ali, M.M., K.K. Sellers, and F. Fröhlich, Transcranial alternating current stimulation modulates large-scale cortical network activity by network resonance. Journal of Neuroscience, 2013. 33(27): p. 11262–11275.

13. Fingelkurts, A.A., et al., Impaired functional connectivity at EEG alpha and theta frequency bands in major depression. Human brain mapping, 2007. 28(3): p. 247–261.

14. Dell’Acqua, C., et al., Increased functional connectivity within alpha and theta frequency bands in dysphoria: A resting-state EEG study. Journal of Affective Disorders, 2021. 281: p. 199–207.

15. Mohammadi, Y. and M.H. Moradi, Prediction of Depression Severity Scores Based on Functional Connectivity and Complexity of the EEG Signal. Clinical EEG and Neuroscience, 2021. 52(1): p. 52–60.

16. Jeong, H.-G., et al., Distinguishing quantitative electroencephalogram findings between adjustment disorder and major depressive disorder. Psychiatry investigation, 2013. 10(1): p. 62.

17. Leuchter, A.F., et al., Resting-state quantitative electroencephalography reveals increased neurophysiologic connectivity in depression. PloS one, 2012. 7(2): p. e32508.

18. Riddle, J., et al., Cross-frequency coupling of neural oscillations in reward-based decision-making dissociates dimensions of depression. submitted, 2021.

19. Riddle, J., A. McFerren, and F. Frohlich, Causal role of cross-frequency coupling in distinct components of cognitive control. Progress in Neurobiology, 2021: p. 102033.

20. Riddle, J., et al., Disinhibition of right inferior frontal gyrus underlies alpha asymmetry in women with low testosterone. Biological Psychology, 2021. 161: p. 108061.

21. Fröhlich, F., Experiments and models of cortical oscillations as a target for noninvasive brain stimulation. Progress in brain research, 2015. 222: p. 41–73.

22. Huang, Y., et al., Realistic volumetric-approach to simulate transcranial electric stimulation—ROAST—a fully automated open-source pipeline. Journal of neural engineering, 2019. 16(5): p. 056006.

23. Lang, P.J., M.M. Bradley, and B.N. Cuthbert, International affective picture system (IAPS): Technical manual and affective ratings. NIMH Center for the Study of Emotion and Attention, 1997. 1: p. 39–58.

24. Voytek, B., et al., Age-related changes in 1/f neural electrophysiological noise. Journal of Neuroscience, 2015. 35(38): p. 13257–13265.

25. Riddle, J., et al., Distinct oscillatory dynamics underlie different components of hierarchical cognitive control. Journal of Neuroscience, 2020. 40(25): p. 4945–4953.

26. Berkman, E.T. and M.D. Lieberman, Approaching the bad and avoiding the good: Lateral prefrontal cortical asymmetry distinguishes between action and valence. Journal of cognitive neuroscience, 2010. 22(9): p. 1970–1979.

27. Harmon‐Jones, E., Clarifying the emotive functions of asymmetrical frontal cortical activity. Psychophysiology, 2003. 40(6): p. 838–848.

28. Jesulola, E., et al., Frontal alpha asymmetry as a pathway to behavioural withdrawal in depression: Research findings and issues. Behavioural Brain Research, 2015. 292: p. 56–67.

29. De Pascalis, V., et al., Relations among EEG-alpha asymmetry, BIS/BAS, and dispositional optimism. Biological Psychology, 2013. 94(1): p. 198–209.

30. Coan, J.A. and J.J. Allen, Frontal EEG asymmetry and the behavioral activation and inhibition systems. Psychophysiology, 2003. 40(1): p. 106–114.

31. Nusslock, R., K. Walden, and E. Harmon-Jones, Asymmetrical frontal cortical activity associated with differential risk for mood and anxiety disorder symptoms: An RDoC perspective. International Journal of Psychophysiology, 2015. 98(2): p. 249–261.

32. Nusslock, R. and L.B. Alloy, Reward processing and mood-related symptoms: An RDoC and translational neuroscience perspective. Journal of Affective Disorders, 2017. 216: p. 3–16.

33. Vrieze, E., et al., Reduced reward learning predicts outcome in major depressive disorder. Biological psychiatry, 2013. 73(7): p. 639–645.

34. Admon, R. and D.A. Pizzagalli, Dysfunctional reward processing in depression. Current Opinion in Psychology, 2015. 4: p. 114–118.

35. Schlaepfer, T.E., et al., Deep brain stimulation to reward circuitry alleviates anhedonia in refractory major depression. Neuropsychopharmacology, 2008. 33(2): p. 368–377.

36. Russo, G.B., et al., Behavioral activation therapy during transcranial magnetic stimulation for major depressive disorder. Journal of affective disorders, 2018. 236: p. 101–104.

37. Lang, P.J. and M.M. Bradley, Emotion and the motivational brain. Biological psychology, 2010. 84(3): p. 437–450.

38. Bergmann, T.O., et al., Combining non-invasive transcranial brain stimulation with neuroimaging and electrophysiology: current approaches and future perspectives. Neuroimage, 2016. 140: p. 4–19.

39. Noury, N. and M. Siegel, Phase properties of transcranial electrical stimulation artifacts in electrophysiological recordings. Neuroimage, 2017. 158: p. 406–416.

40. Hanslmayr, S., J. Matuschek, and M.-C. Fellner, Entrainment of prefrontal beta oscillations induces an endogenous echo and impairs memory formation. Current biology, 2014. 24(8): p. 904–909.

41. Wang, J.X., et al., Targeted enhancement of cortical-hippocampal brain networks and associative memory. Science, 2014. 345(6200): p. 1054–1057.

42. Riddle, J., D.R. Rubinow, and F. Frohlich, A case study of weekly tACS for the treatment of major depressive disorder. Brain Stimulation: Basic, Translational, and Clinical Research in Neuromodulation, 2020. 13(3): p. 576–577.

