## Supplemental Material for "Reduction in left frontal alpha oscillations by transcranial alternating current stimulation in major depressive disorder is context-dependent in a randomized-clinical trial"

Supplemental Material for Riddle et al.

*Sections of the Supplemental Material are presented in order of appearance in the manuscript.*

---

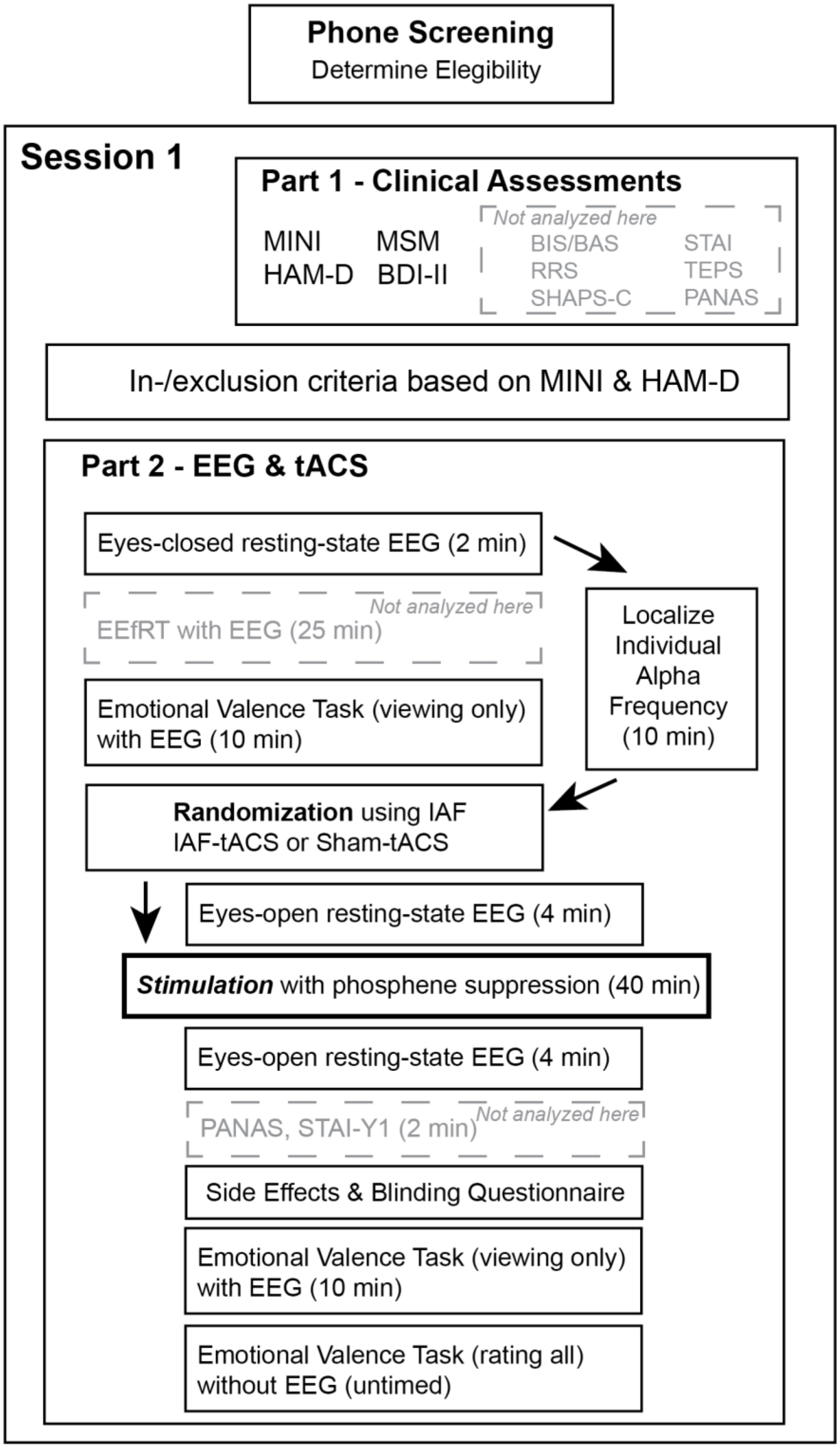


**Supplemental Figure 1**. *Flow chart for the experiment*. Participants that passed the phone screening were invited to participate in the main experiment. The first and only in-person session comprised two parts. In the first part, clinical assessments were administered. For this manuscript, only a subset of assessments related to depression severity and major depressive disorder diagnosis were analyzed. Other assessments were included in another analysis that is not published here. After part 1, participants were screened for eligibility (see inclusion/exclusion criteria). In part 2 of the session, an eyes-open resting-state EEG was collected and these data were analyzed to extract the individual alpha frequency from parietal-occipital electrodes. This analysis was conducted while participants performed two tasks: the Expenditure of Effort for Reward Task (EEfRT) that was not analyzed for the sake of this manuscript, and the emotional valence task. At this time, participants passively viewed images from the international affective picture system (IAPS) for two seconds each. Note that the participants did not rate the valence of the images at this time. Participants completed the four-minute pre-stimulation eyes-open resting-state EEG. Next, participants were randomized to either IAF-tACS or to sham-tACS and a code was selected that corresponded to their IAF from 8 to 13 Hz in 0.5 Hz increments. Then, participants completed 40 minutes of stimulation during viewing of a wide-angle underwater scene for phosphene suppression. Participants completed the post-stimulation four-minute eyes-open resting-state EEG. Participants completed two clinical assessments that were not analyzed for this manuscript, and completed the side-effects and blinding questionnaire. Then, participants completed the emotional valence task with passive viewing of novel images. Finally, participants rated all of the images that they had seen before and after stimulation at their own pace without EEG recording. MINI is the Mini International Neuropsychiatric Interview for the DSM-V. MSM is the Maudsley Staging Method. HAM-D is the Hamilton depression rating scale. BDI-II is the Beck's Depression Inventory version 2. BIS/BAS is the Behavioral Inhibition System and Behavioral Approach System scale. STAI is the state-trait anxiety inventory. TEPS is the temporal expectation of pleasure scale. RRS is the ruminative responses scale. PANAS is the positive and negative affect schedule. SHAPS-C is the Snaith-Hamilton pleasure scale clinician administered.

---

**Supplemental Section 1**: *Inclusion and exclusion criteria*

The exclusion criteria were as follows: suicide risk as determined by the Mini-International Neuropsychiatric Interview (MINI) for the DSM-5 [1] and three or greater on the suicidality metric of the Hamilton Depression Rating Scale (HAM-D) [2], neurological illness, history of traumatic brain injury, prior brain surgery, any implanted devices, pregnant or nursing females, current use of benzodiazepines or anti-epileptic drugs, non-English speaking, MINI diagnosis of substance use disorder within the last 12 months, substance use within the last 12 months (other than nicotine or cannabis, but note that participants were required to pass a urine drug test on the day of the experiment), eating disorder (current or within the past three months). For participants that reported to experience depression in episodes (e.g. bipolar depression), those that were not in a depressive episode were excluded. Participants were not excluded on the basis of concomitant psychotherapy or medication use (except those that have robust effect on the EEG, see above). The inclusion criteria for participants with depression were a diagnosis of MDD using the MINI and a HAM-D score of at least 8. For healthy controls, additional exclusion criteria were a current or history of psychiatric illness, and first degree relative with psychiatric illness. 15 participants did not show for their session or discontinued participation before their in-person session. 24 participants were determined to be ineligible: six participants reported high suicide risk and an acute psychological assessment was conducted, six participants did not meet our depression threshold, five participants with MDD were excluded (current eating disorder, two substance use disorder, receiving transcranial magnetic stimulation, hypomanic episode), four healthy control participants did not meet criteria, and three participants tested positive for THC. After determining eligibility, 87 participants began the experiment. The experiment was discontinued for three participants during the baseline task, before randomization, because one participant fell asleep during the task, one participant experienced mild anxiety, and one participant left the experiment due to unrelated nausea. Thus, 84 participants were randomized to one arm of the study (verum or sham) such that an age and sex matched pair of participants received the same form of stimulation and the total number receiving each was balanced. After randomization, a technical difficulty occurred for one participant and that participant did not receive the allocated intervention and participation was discontinued. Finally, one participant was difficult to age and sex match; thus, they were excluded from the analysis. The final count of participants that completed the study and were analyzed was 82 (66 women): 41 healthy controls and 41 participants with MDD.

---

**Supplemental Section 2**: *EEG Preprocessing Pipeline*

Both datasets, resting-state and emotional valence task, were preprocessed using the EEGLAB toolbox in MATLAB [3], and each dataset was preprocessed separately such that artifacts unique to each recording could be isolated. In addition, the epochs before and after stimulation were concatenated into a single dataset such that differences in preprocessing cannot explain any effect of stimulation. Finally, the experimenter was blind to the stimulation condition of each participant during preprocessing. We applied a high pass filter of 1 Hz and a low pass filter at 58 Hz. Thus, 58 Hz set the upper boundary for analysis of gamma frequency activity (35-58 Hz). Data were downsampled from 1000 Hz to 200 Hz. Data from the emotional image viewing were epoched from 0.9 prior and 2 seconds after the presentation of each image. Data from resting-state was epoched into 2 second periods. Data were then manually inspected and trials corrupted with noise were rejected from future analysis. The data were then cleaned using an artifact subspace reconstruction algorithm to remove high-variance signal and reconstruct missing data [4]. This algorithm also flagged noisy channels, which were then replaced with a spherical interpolation from its neighboring channels. Global average re-referencing was applied, which is an approximate solution for the spherical electrical field assumptions that was enabled by use of a 128-channel system that include electrode coverage on the face and neck. Principal component analysis was run based on the rank of the data matrix to optimize the data for artifact rejection using info-max independent component analysis. All independent components were visually inspected, and components that corresponded to line noise, muscle activity, eye movement, blinks, and heart beat were removed from the data.

---

**Supplemental Section 3**: *Individual Alpha Frequency Localization*

Individual alpha frequency was calculated during the experiment as the participant performed two tasks. An abbreviated preprpocessing pipeline was used. The two-minutes of rest were extracted, band-pass filter was applied, and data was re-referenced to the global average. Then, the fast Fourier transform was applied to the entire timeseries, the average power spectra of parietal-occipital electrodes (P7, Oz, P8, P4, Pz, and P3 comprised the perimeter) was calculated, and the maximum frequency within the alpha band (8-13 Hz) was extracted. After visual inspection of the resulting power spectrum and IAF, the resulting value in 0.5 Hz increments was defined as IAF. If an alpha peak could not be identified, then the raw data was checked for large artifacts and the preprocessing was re-run after removing that data. If a peak could still not be identified, then a default of 10 Hz was used. Using this method, IAF could not be identified in 6 participants of the 82 participants in the final dataset (four healthy controls). For display purposes in Figure 1A, the analysis was repeated with more extensive preprocessing (see previous description sans ICA), the power spectra of each 4 second epoch was calculated and the median power spectrum was calculated for each channel, then the spectra were averaged (mean) in the parietal-occipital electrodes. Using the more extensive preprocessing pipeline, IAF could not be identified in only two participants, both of whom were healthy controls. The mean IAF using advanced preprocessing was similar to IAF derived from simple preprocessing (10.2±0.9 Hertz in patients with MDD and 10.1±0.8 Hertz in healthy controls). As an exploratory analysis, we calculated IAF in the left frontal electrodes (F3 and surrounding) and compared the peak frequency (9.9±0.9 in patients with MDD and 9.9±0.9 in healthy controls). We found that IAF in left frontal electrodes could not be identified in four participants. There was a systematic decrease in IAF for the left frontal electrodes relative to the parietal-occipital electrodes (pair-wise Student's t-test, N = 78, t(77) = 3.623, p = 0.00052, d = 0.410). The average for left frontal electrodes was 9.914 Hz with a standard deviation of 0.9.06. The average difference in IAF between left frontal electrodes and parietal-occipital electrodes, while systematic, was less than the resolution of 0.5 Hz that was used for the stimulation frequency. Thus, it is unlikely that this difference made a difference in practice. In addition, we theorize that the alpha frequency is a robust carrier frequency for inter-regional communication and, thus, general consistency between brain regions in IAF is expected. Furthermore, alpha oscillations are implicated in thalamo-cortical interactions and thus a common driver in thalamus may support the similarity in IAF.

---

**Supplemental Section 4:** *Double-blind Placebo-Control IAF-tACS*

Stimulation was delivered using three silicone-carbon electrodes attached to the scalp using electrically conductive paste (Ten20, Bio-Medical Instruments, Clinton Township, MI, USA). Target electrodes, 5x5 centimeters, were centered on F3 and F4 with a split wire such that both frontal cortices received synchronized electrical input. The return electrode, 5x7 centimeters, was oriented posterior to anterior and centered on Cz. Electrode placement was determined using scalp measurements according to the international 10-20 measurement system. A 1-centimeter diameter circle was cut out of the return electrode such that the reference electrode (Cz) on the EEG net was directly on the scalp and was not bridged with the stimulation electrode. Impedance was ensured to be below 10 kΩ before stimulation began. Six-digit numeric codes were generated for each participant for every possible IAF, and the mapping between stimulation condition (verum or sham) was kept hidden from experimenters and maintained by a lab member not involved in the research. To ensure accurate stimulation, the tACS device recorded and encrypted waveforms from stimulation. These waveforms were checked after each experimental session by a researcher not involved with the experiment. Verum stimulation consisted of 40 minutes of alternating current at 1 mA (zero-to-peak) delivered to each frontal target electrode and 2 mA received from the return electrode. We utilized an active sham. Participants from the same population in our previous experiment were successfully blinded with the same active sham [5]. Active sham consisted of a 20 second ramp up 40 seconds of stimulation and then a 20 seconds ramp down. This stimulation mimics the typical skin sensation at the onset of stimulation. Experimenters and participants completed a blinding questionnaire after stimulation was complete. A common side effect of tACS is the illusory perception of flickering lights, called phosphenes, that can reduce the ability to blind participants to which session is sham [6]. During stimulation, participants watched a colorful video with frequent motion and luminance changes to reduce phosphene perception (Reefscapes, Undersea Productions, Queensland, Australia). The video consists of wide-angle, long-form recordings of actual coral reefs with fish and other wildlife. After stimulation, the tACS device was unplugged from the electrodes during all EEG recordings to ensure that there was no coupling between the EEG sensors and the battery-powered electronics of the device. When the battery was removed from the device it was placed in a bin of used batteries. These used batteries were periodically charged in a separate room from the experimental sessions. This procedure masked the degree to which each battery was drained, because verum stimulation will drain the battery more than sham.

---

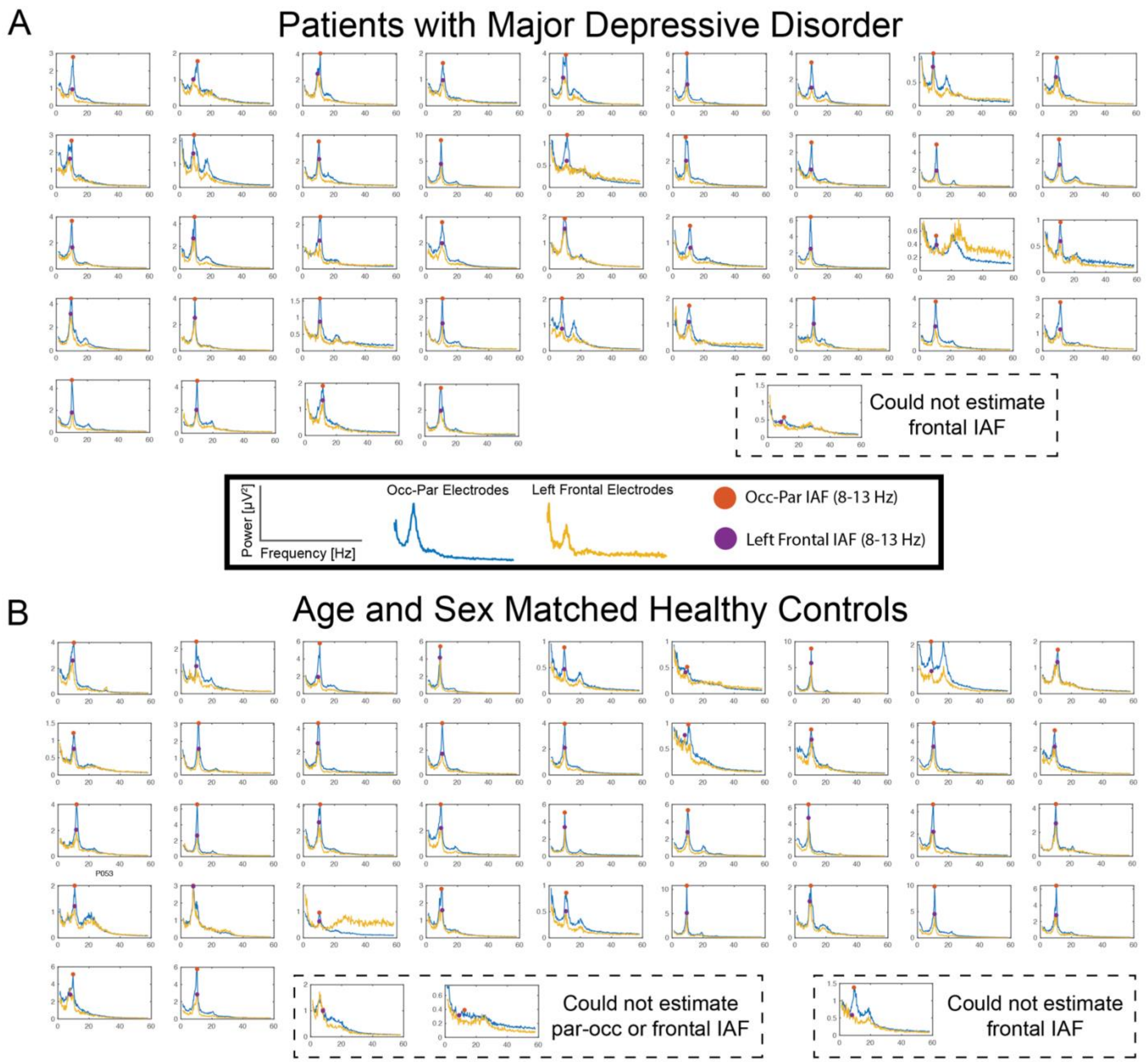


**Supplemental Figure 2**. *Comprehensive assessment of individual alpha frequency localization.* Individual alpha frequency (IAF) was estimated from parietal-occipital electrodes as the frequency with peak alpha power from the range of 8 to 13 Hertz to be used for our primary analysis of the effect of tACS on eyes-open resting-state EEG. Of the 82 participants, IAF for two of the healthy control participants could not be estimated (dashed outline) from the parietal-occipital electrodes after advanced preprocessing. For two additional participants (one participant with MDD and one healthy control), IAF from the left frontal electrodes (F3 and surrounding) could not be estimated. There was a systematic decrease in IAF for the left frontal electrodes relative to the parietal-occipital electrodes (pair-wise Student's t-test, $N = 78$, $t(77) = 3.623$, $p = 0.00052$, $d = 0.410$). The average for left frontal electrodes was 10.1506 Hz with a standard deviation of 0.8251. The average for left frontal electrodes was 9.914 Hz with a standard deviation of 0.9.06.

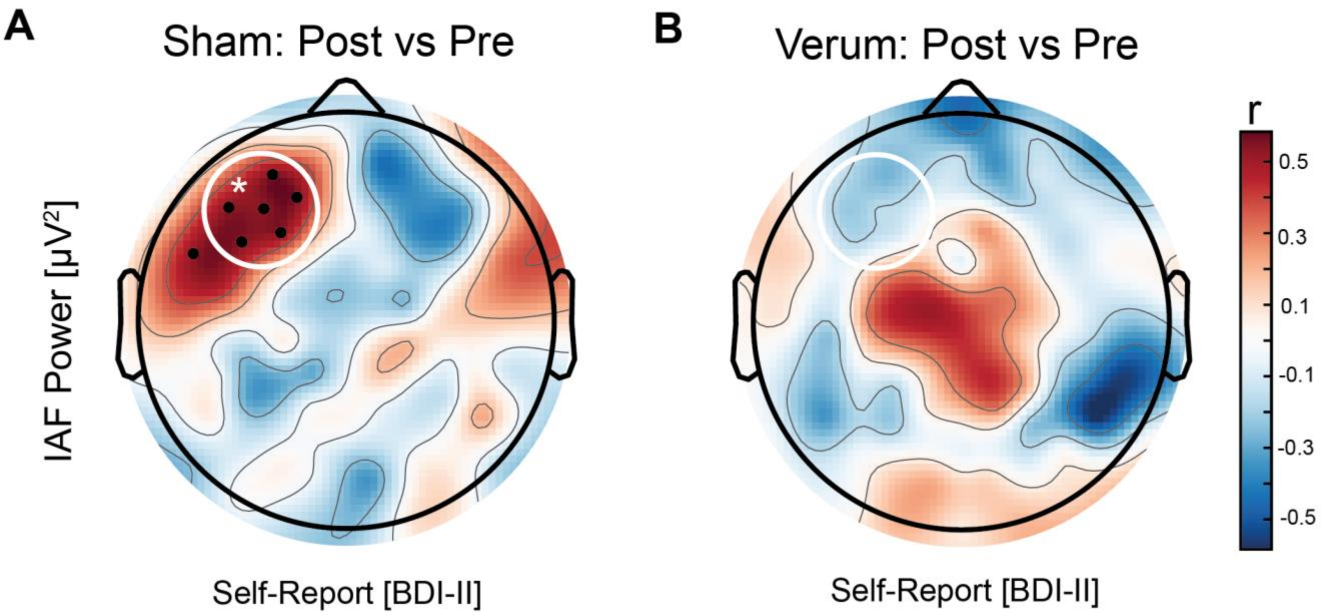


**Supplemental Figure 3**. *Correlation of BDI-II to change in left frontal alpha power to IAF*-tACS. (A) Participants with MDD with the greatest depression severity showed the greatest increase in left frontal IAF power with sham stimulation (N=20, r(19) = 0.512, p = 0.021). The effect was specific to left frontal electrodes (white circle). (B) The relationship between depression severity and left frontal IAF power was negative, but not significant, for verum stimulation (N=21, r(20) = -0.172, p = 0.455) with no significant electrode clusters across the scalp. MI is modulation index. *p<0.05; Units for IAF power were modulation index.

---

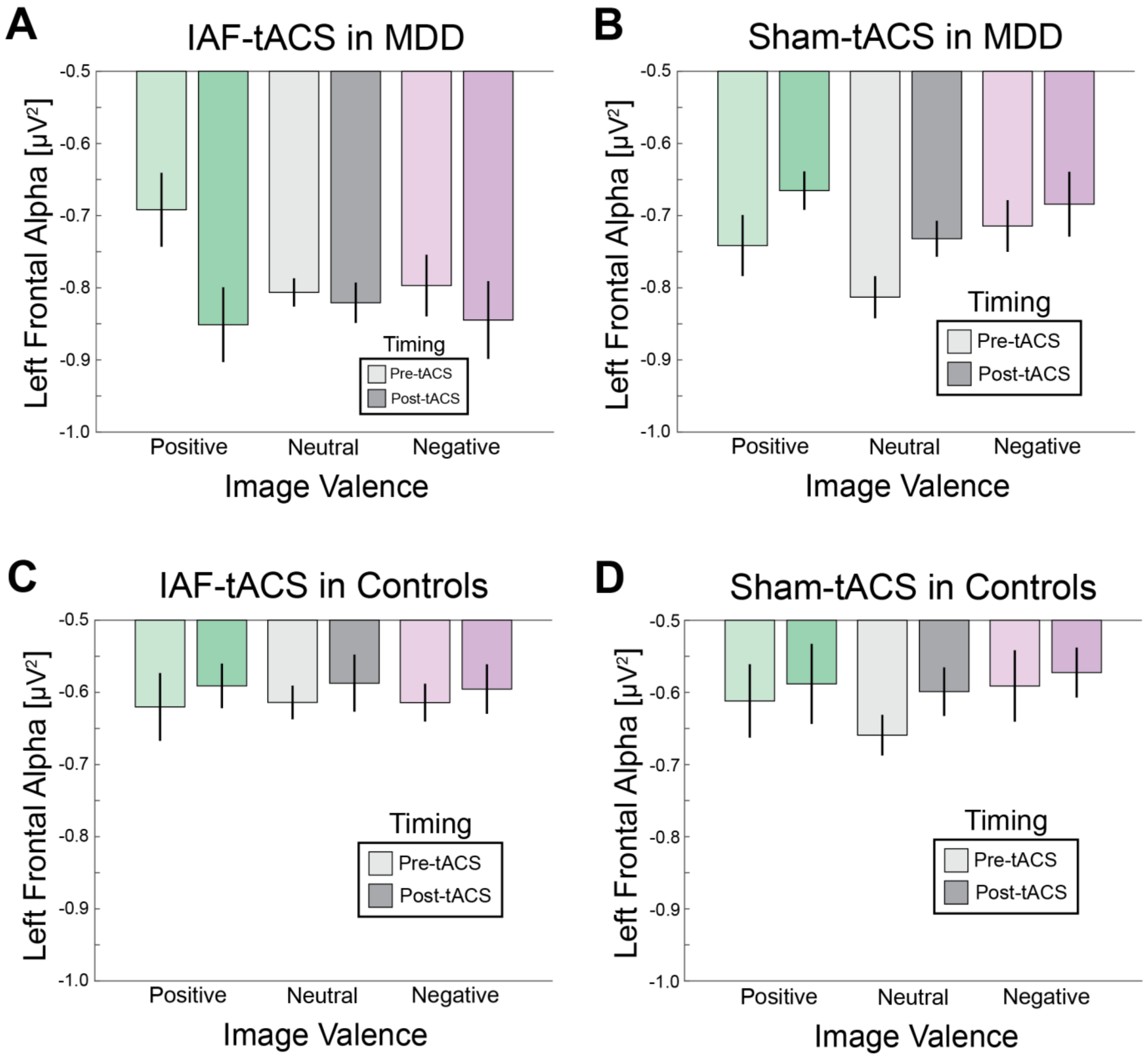


**Supplemental Figure 4**. *Left frontal alpha IAF-tACS effect for all conditions in the emotional valence task*. (A-D) All conditions for the emotional valence task by group (patients with MDD or healthy controls) and stimulation type (IAF-tACS or sham-tACS) are depicted. Positive valence conditions are in green, neutral in grey, and negative in purple. The darker shades depict the post-tACS recording and the lighter shades depict pre-tACS. Data were normalized for spatial topography using the z-transformation across all scalp electrodes. Error bars are SEM.

---

**Supplemental Section 5**: *Comprehensive analysis of variance*

When a two-way ANOVA was performed for the modulation index of left frontal alpha power with factors stimulation (IAF-tACS or sham tACS) and depression severity (HAM-D as a continuous variable) for all participants, we found a trend-level interaction between depression severity and stimulation (N=82, $F(1,78)=3.128$, $p=0.081$, $\eta_p^2=0.04$). This analysis recapitulated the overall pattern of findings, in which the difference between verum and sham was present only in those with MDD and greater depression severity. This analysis also revealed a trend-level main effect of stimulation ($F(1,78)=3.389$, $p=0.069$, $\eta_p^2=0.04$) such that left frontal alpha power was reduced for verum stimulation relative to sham most likely driven by the strong effect in participants with MDD. There was no main effect of depression severity irrespective of stimulation type ($F(1,78)=0.038$, $p=0.846$, $\eta_p^2=0.00$).

**Supplemental References**


1. Sheehan, D., et al., *The validity of the Mini International Neuropsychiatric Interview (MINI) according to the SCID-P and its reliability.* European psychiatry, 1997. **12**(5): p. 232-241.
2. Williams, J.B., *A structured interview guide for the Hamilton Depression Rating Scale.* Archives of general psychiatry, 1988. **45**(8): p. 742-747.
3. Delorme, A. and S. Makeig, *EEGLAB: an open source toolbox for analysis of single-trial EEG dynamics including independent component analysis.* Journal of neuroscience methods, 2004. **134**(1): p. 9-21.
4. Mullen, T., et al., *Real-time modeling and 3D visualization of source dynamics and connectivity using wearable EEG.* Conference proceedings : ... Annual International Conference of the IEEE Engineering in Medicine and Biology Society. IEEE Engineering in Medicine and Biology Society. Annual Conference, 2013. **2013**: p. 2184-7.
5. Alexander, M.L., et al., *Double-blind, randomized pilot clinical trial targeting alpha oscillations with transcranial alternating current stimulation (tACS) for the treatment of major depressive disorder (MDD).* Translational psychiatry, 2019. **9**(1): p. 1-12.
6. Kar, K. and B. Krekelberg, *Transcranial electrical stimulation over visual cortex evokes phosphenes with a retinal origin.* Journal of Neurophysiology, 2012. **108**(8): p. 2173-2178.
